# Shopping with glaucoma. Quantifying the impact of mild glaucomatous visual field loss using a virtual reality supermarket

**DOI:** 10.64898/2026.01.26.26344620

**Authors:** Peter F. Reddingius, David P. Crabb, Pete R. Jones

**Author notes:** **<u>CORRESPONDENCE</u>** Pete R Jones, Department of Optometry and Visual Science; School of Health and Medical Sciences; City St George’s, University of London; Northampton Square; London; UK; EC1V 0HB.

## Abstract

Conventional assays (questionnaires, acuity, visual fields) may be insufficient to assess the real-world impact of new glaucoma treatments. Here, we report a novel virtual reality shopping task, and assess its sensitivity to differences in mild visual field loss. Eight glaucoma patients were asked to freely navigate a virtual grocery store and place items from a shopping list into a trolley. Across a range of metrics (time, head/body movements), performance was associated with variations in binocular visual field loss [*r*_238_=0.35, *P*<.001]. This indicates promise for using virtual reality tasks to evaluate the benefits of new treatments. Improvements and use-cases are discussed.

**ONLINE ONLY SUPPLEMENTARY MATERIAL:** The article contains supplementary material which will be provided as a single stand-alone PDF document.

## I. INTRODUCTION

To determine whether new treatments offer good value, we need to quantify how they improve patients’ everyday quality of life (QoL). Typically, this is achieved using Patient Reported Outcome Measures (PROMs). However, PROMs alone may be insufficient, owing to their subjective nature, inherent noise,[1] and practical limitations.[2] We have therefore been exploring whether it is possible to complement PROMs using direct measures of real-world functional vision, designed to give convergent information about a patient’s ability to perform everyday tasks.

Here, we report a novel virtual reality shopping task (an everyday task that many glaucoma patients report difficulties with[3]). We assessed its feasibility in eight glaucoma patients, and measured its sensitivity to variations in mild binocular VF loss. We specifically selected individuals with mild binocular loss as this is the group where conventional PROMs are thought to be least sensitive,[4] yet also where preventative interventions are likely to be most beneficial.[5]

## II. METHODS

### 2.1 Experimental setup

Participants wore a virtual reality headset weighing 722g (Meta Quest Pro; Meta; USA) and physically walked around a virtual reality supermarket environment with no controllers or trackers attached. As shown in **Figure 1A**, participants were given a virtual shopping list of items to find (30 items on the list, spread over 6 aisles), and were required to place found items into a shopping trolley using a reach-and-grasp motion (optical hand tracking; see **Figure 1B&C**). The task was completed binocularly (no patching), and in general every effort was made to make the task as “true to life” as possible. Items varied naturally in visibility depending on their location (e.g., items on lower shelves were less well illuminated), the legibility of their label (e.g., contrast, font size), and how many other similar items were present (e.g., there were three “brands” of crisps with ∼5 flavours each). When placing items into the trolley the participants were given feedback as to whether or not that item was correct (i.e., was on the shopping list), and the task continued until all items on the list were successfully deposited into the trolley. For further technical details, see **Supplementary Section 1**. For an example screen recording see https://youtu.be/mTwC5Xjcpqs.

**Figure 1.**
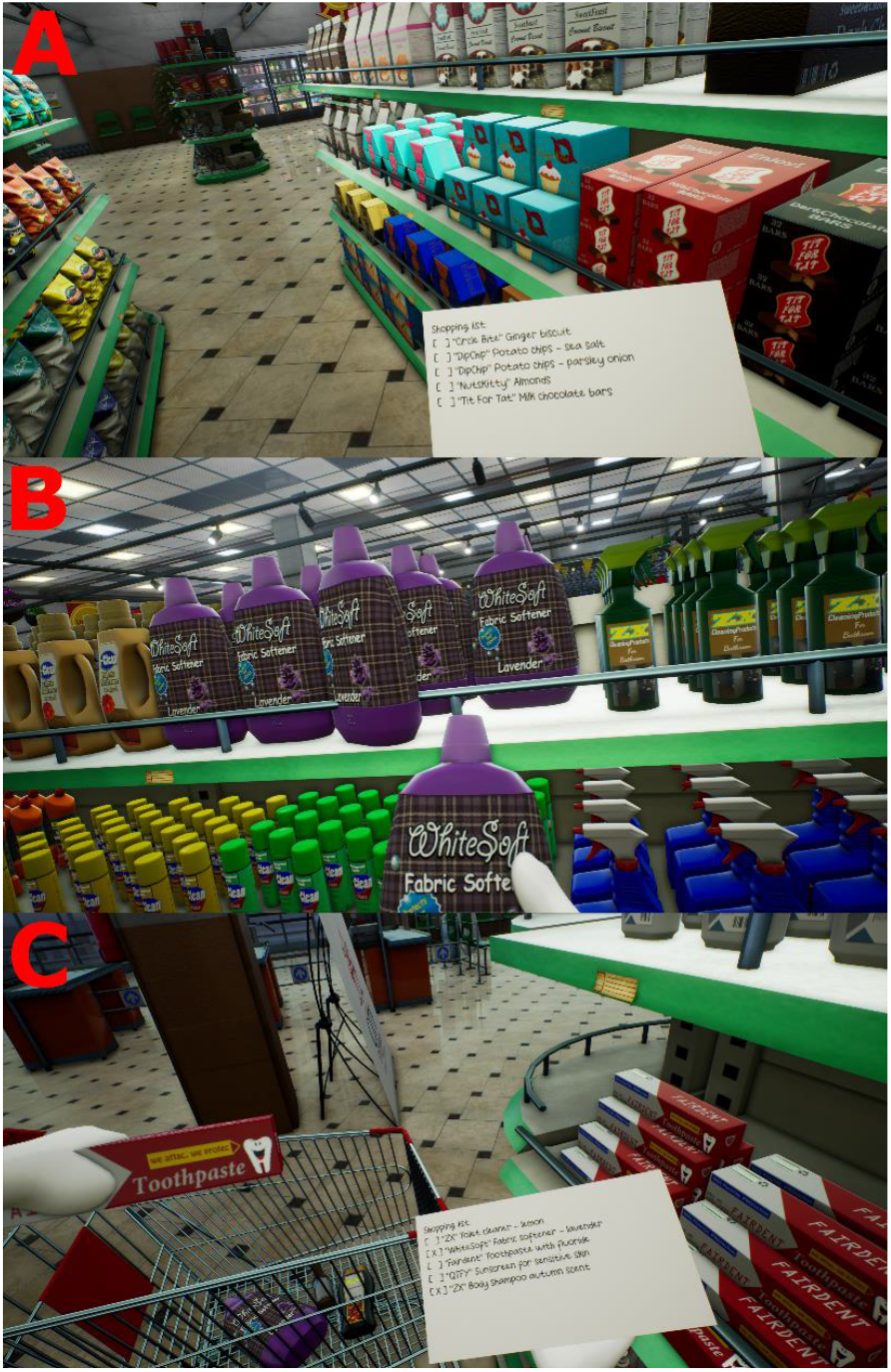
(**A**) A person wearing the VR headset and looking at the shopping list while in the aisle. (**B**) The person reaching for an item on the shelf. (**C**) The person holding an item above the shopping trolley while checking the shopping list. Some items have been deposited into the trolley already and are marked with an “X” on the list. Images are screenshots of the internal view of the headset.

### 2.2 Participants

Participants consisted of eight glaucoma patients (*see* **Supplementary Table 1** for details), whose diagnoses were confirmed by an ophthalmologist. All participants underwent monocular VF assessments (Humphrey Field Analyser v3.0, SITA Fast), and all of their fields can be found in **Supplementary Figure 1**.

For each participant, we calculated their expected binocular VF using the integrated VF (IVF) method (taking the best pointwise value for each eye).[6] Note that as shown in **Figure 2**, some participants exhibited moderate to advanced monocular VF damage (e.g., G3, G6), but the expected binocular IVF loss of all patients was still mild (< -6dB).[7]

**Figure 2.**
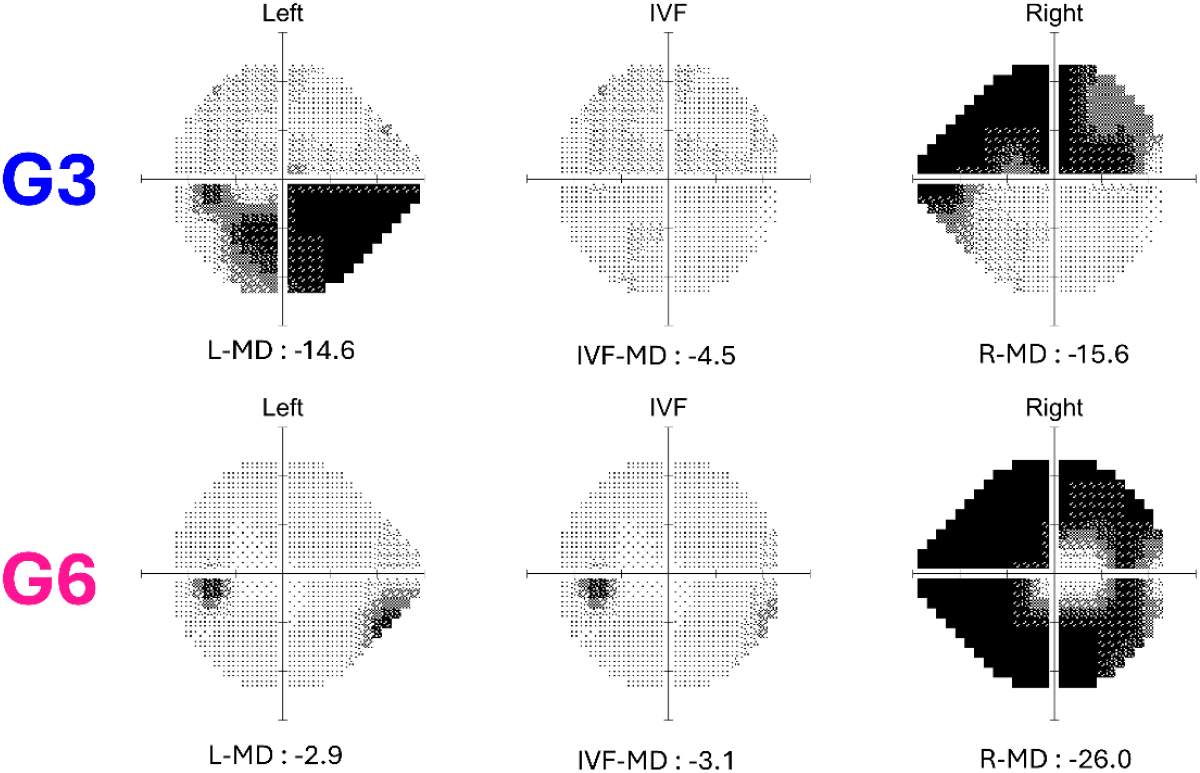
Images of the visual field for two glaucoma patients (G3 and G6). The left and right monocular visual fields were measured empirically using the Humphrey Field Analyzer (ZEISS; Oberkochen, Germany). The integrated visual field (IVF) plots show the predicted binocular visual fields, when taking the maximum of the left/right eye pointwise values.[6] Numbers show the Mean Deviation values, computed using the control data from the visualFields R package.[8]

### 2.3 Analysis

Aside from computing time taken and number of errors, we also computed various movement metrics using the VR headset sensors (e.g., hand, eyes, and head movements). To avoid confounding total movement with time taken, movement vectors were first summed and then divided by time.

Primary analyses consisted of Pearson’s *r* correlations. Pearson’s was preferred for ease of interpretation and comparison with other studies. However, since normality was violated, all reported results were also confirmed using a non-parametric Kendall tau test, which produced qualitatively similar results (no change in significance/non-significance, and significant *P* values generally agreeing to three decimal places).

## III. RESULTS

The principal finding was that variations in mild VF loss (as measured by the IVF) were correlated with overall performance on the task [*r*_238_=0.35, *P*<0.001], see **Figure 3**.

**Figure 3.**
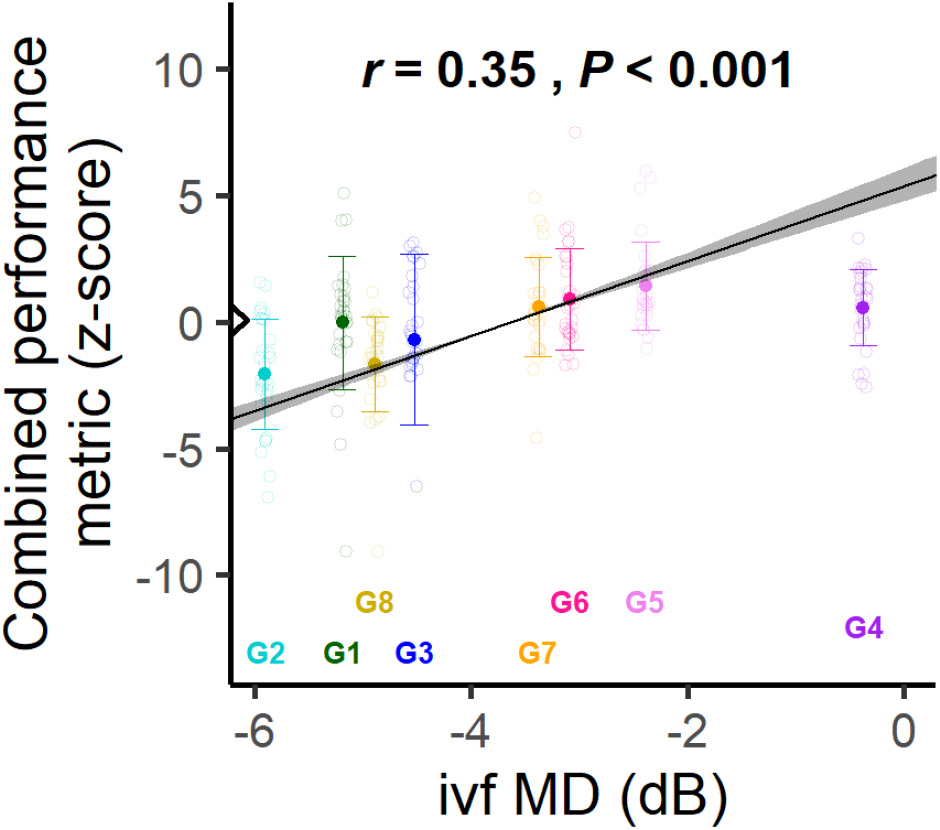
Association between magnitude of visual field loss (IVF-MD) and task performance. The combined performance metric is a summation of z-scores of four individual metrics described in the results section. The IVF-MD is the mean deviation of the integrated visual field.[6] Error bars indicate mean ± standard deviation for each of the 8 glaucoma patients (represented with their ID and colour, see **Supplementary Table 1**). For comparison, the triangle on the y-axis shows the grand mean value for 9 control participants (median {IQR} age = 68 {65-74} years). Unfilled circles show the task performance for each item the glaucoma patients collected. The black line signifies the standard major axis regression, with shaded regions indicating the slope’s 95% confidence interval. Black text at the top signifies Pearson’s correlation coefficient and significance value.

Note that this overall performance metric was computed by combining several individual variables (see **Supplementary Figure 2**), and many of these individual variables were also associated with VF loss. Specifically: patients with greater VF loss moved their head less [*r*_238_=0.29, *P*<0.001], translated their body less [*r*_238_=0.29, *P*<0.001], made more grabbing movements for target items (e.g., because they dropped it, or were hesitant or uncertain) [*r*_238_=-0.17, *P*=0.008], and spent longer moving items from the shelf to the shopping trolley [*r*_238_=-0.20, *P*=0.002].

In contrast to binocular VF loss, performance was not correlated with the worse eye’s MD [*r*_238_=-0.01, *P*=.865]. And there were no significant associations between binocular (IVF) loss and time to find items [*r*_238_=-0.09, *P*=0.149], time spent reading the shopping list [*r*_238_=0.07, *P*=0.298], or the number of incorrect items that were put into the trolley [*r*_238_=-0.03, *P*=0.600].

## IV. DISCUSSION

This initial proof-of-principle study demonstrates that it is possible to use virtual reality to objectively measure variations in performance on a meaningful real-world task.

As binocular VF loss increased, people with glaucoma exhibited reduced body translation, took longer walking to the shopping trolley, made more head movements, and made more reaching movements. All of these differences could be summarised in a single z score, which in the future could potentially be used to track progression or assess treatment efficacy.

The present findings are broadly consistent with previous studies. For example, Lam et al. (2020)[9] showed that the binocular VF of glaucoma patients was correlated with the time to find items in a virtual reality grocery store. That is partially consistent with our findings, as our combined metric was correlated with IVF-MD, however, we saw no correlation between search time specifically and IVF-MD. This could be because in Lam’s study most glaucomatous eyes were of moderate to advanced glaucoma. We speculate that additional deficits might also be observed using the present task if participants with moderate-to-severe vision loss were included. However, we purposefully excluded these in the current study to evaluate if a ‘signal’ could be found in mild binocular VF loss.

Although the association was significant, the effect size of VF loss was relatively small [*r*=0.35], and there was considerable individual variability (see **Figure 3**). This is, to some extent, inevitable given the complex and unrestricted nature of using a “real-world” task. However, it may be that the effect size could be increased by introducing additional elements that people with glaucoma report difficulties with, such as reducing lighting or low contrast labels.[10] It also remains to be seen whether some of this individual variability can be mitigated by using this technology to longitudinally follow the same individuals over time (e.g., within-subject analyses, before-and-after treatment).

In short, the present study demonstrates that mild binocular glaucomatous visual field loss is correlated with reduced performance in the virtual reality shopping task. Next steps (currently ongoing) include assessing the patient acceptability of such VR assessments, and standardising/optimising the testing procedure for potential use in clinical trials.

## Data Availability

All data produced in the present study are available upon reasonable request to the authors

## ACKNOWLEDGEMENTS

We thank Dr. Jeremy Tan for confirming the diagnosis of the glaucoma patients. We additionally thank Dr. Jeremy Tan, Dr. Aiman Hafeez and Sajni Bohra for their help recruiting participants and conducting vision tests. This research was supported by the European Union’s Horizon 2020 research and innovation programme under the Marie Sklodowska-Curie Grant Agreement No. 955590.

## AUTHORSHIP CONTRIBUTIONS

**PFR**: Conceptualisation; Methodology; Software; Investigation; Data curation; Formal analysis; Writing – original draft.

**DPC**: Conceptualisation; Methodology; Writing – review and editing; Supervision.

**PRJ**: Conceptualisation; Methodology; Writing – review and editing; Supervision

