## Supplementary material for "Shopping with glaucoma. Quantifying the impact of mild glaucomatous visual field loss using a virtual reality supermarket"

**Manuscript title**

### Supplementary Section 1: Technical details

Participants wore a Meta Quest Pro (Meta, Menlo Park, USA) headset which contains two 2k (1800x1920 pixels) monitors running at 70Hz and has a combined field of view of 106x96°. The headset was connected to a PC (Dell, Round Rock, USA) with a GTX4080 graphics card (Nvidia, Santa Clara, USA) using a Meta Link Cable (Meta, Menlo Park, USA) and a 5m extension cable (5m USB 3.2 5Gbps C/C active extension; Lindy Electronics, Thornaby, UK).

The virtual environment was running in the Oculus branch of Unreal Engine 4.27 (Epic Games, Cary, USA). Before starting the experiment, participants spent ~20 minutes (until they reported they felt comfortable) in a training environment to get accustomed to the way the task worked and to reduce learning effects. The training environment was highly similar to the test environment except there were fewer items to not overwhelm participants.

### Supplementary Section 2: Participant details

See **Supplementary Table 1** for all demographic details of the participants and see **Supplementary Figure 1** for the greyscales of the patient's visual fields including their integrated visual field.

| ID | Age range | diagnosis | Sex | OS MD | OD MD | IVF MD | VA OS | VA OD |
|---|---|---|---|---|---|---|---|---|
| **G1** | 60-65 | POAG | F | -12.16 | -7.32 | -5.19 | 0 | 0.2 |
| **G2** | 70-75 | POAG | M | -9.27 | -10.76 | -5.90 | 0.1 | 0.2 |
| **G3** | 60-65 | POAG | F | -14.63 | -15.59 | -4.52 | 0.7 | 0.2 |
| **G4** | 50-55 | POAG | M | -2.85 | -0.46 | -0.37 | 0 | 0.2 |
| **G5** | 70-75 | POAG | M | -2.93 | -3.45 | -2.39 | 0.1 | 0.4 |
| **G6** | 60-65 | POAG | M | -2.89 | -25.99 | -3.09 | 0.3 | 0 |
| **G7** | 60-65 | SOAG | M | -4.08 | -14.68 | -3.38 | 0 | 0.3 |
| **G8** | 70-75 | ACG | F | -8.9 | -9.09 | -4.89 | 0.1 | 0.2 |

***Supplementary Table 1.*** *Demographic information of the glaucoma patients.*

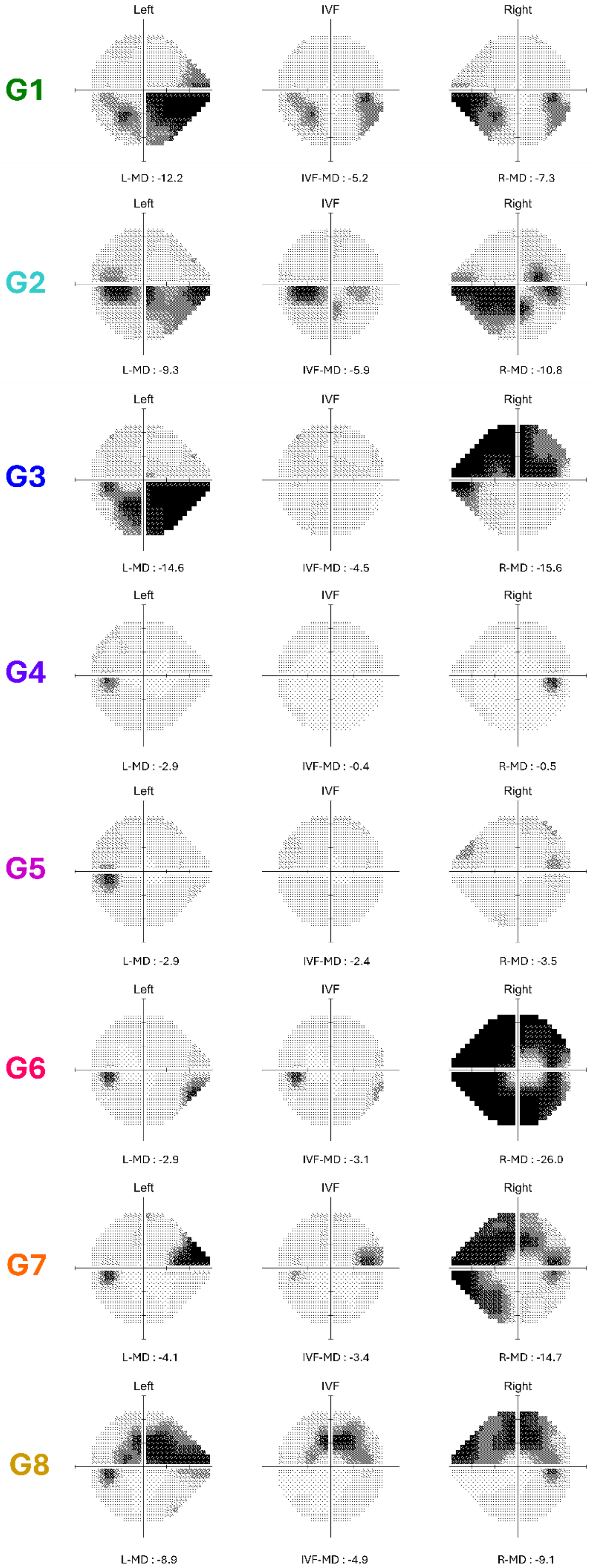
Left
IVF
Right
G1
L-MD : -12.2
IVF-MD : -5.2
R-MD : -7.3
G2
L-MD : -9.3
IVF-MD : -5.9
R-MD : -10.8
G3
L-MD : -14.6
IVF-MD : -4.5
R-MD : -15.6
G4
L-MD : -2.9
IVF-MD : -0.4
R-MD : -0.5
G5
L-MD : -2.9
IVF-MD : -2.4
R-MD : -3.5
G6
L-MD : -2.9
IVF-MD : -3.1
R-MD : -26.0
G7
L-MD : -4.1
IVF-MD : -3.4
R-MD : -14.7
G8
L-MD : -8.9
IVF-MD : -4.9
R-MD : -9.1

***Supplementary Figure 1.*** *Greyscales showing differential light sensitivities across the 24-2 visual field (generated from the raw pointwise data using visualFields R package for control data and MATLAB), for the left eye (left column), right eye (right column), and the predicted integrated visual field (central column) of the eight glaucoma patients.*

**Supplementary Section 3: Additional result figures**

This section shows additional figures of the results mentioned in the main text.

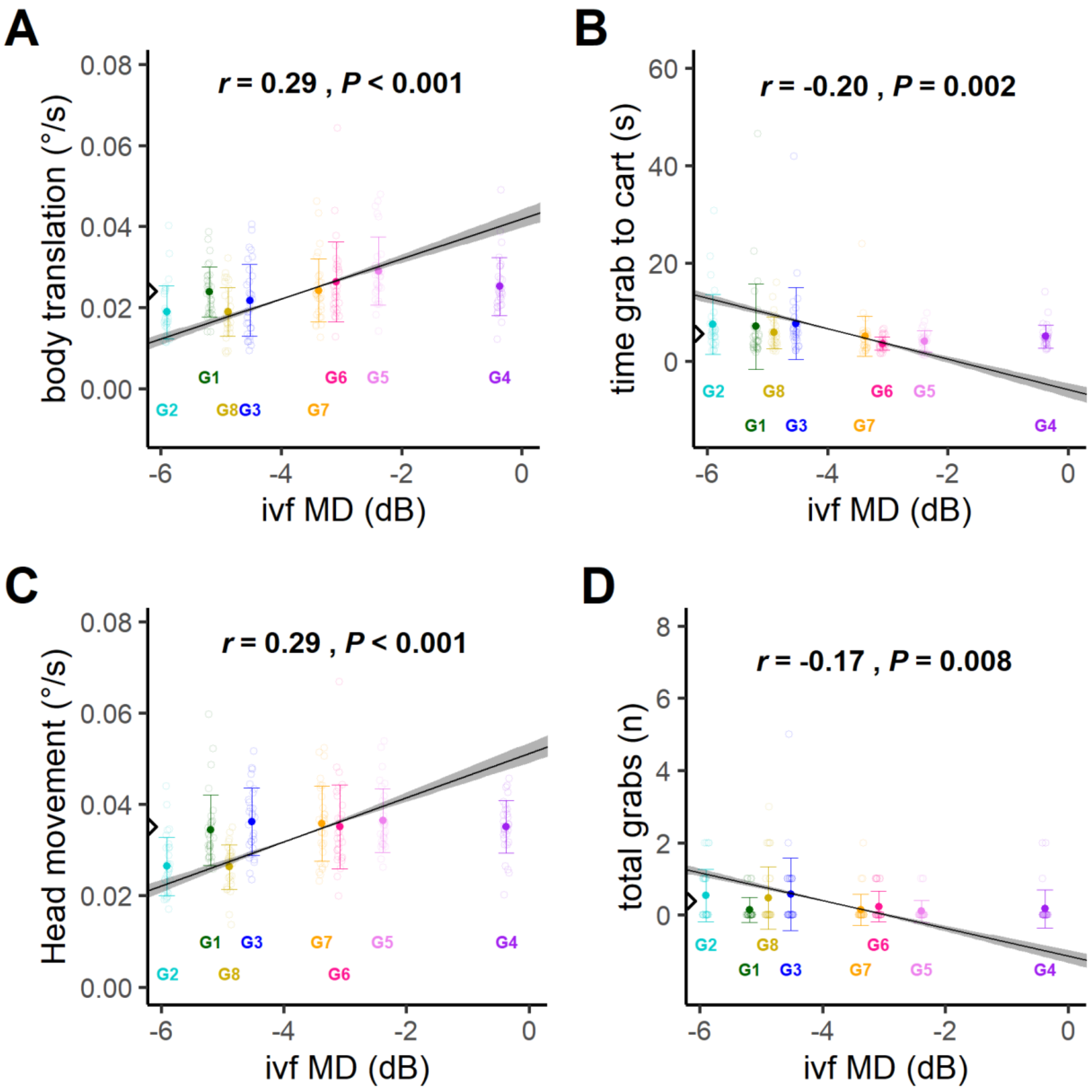


***Supplementary Figure 2.*** *Association between magnitude of visual field loss (IVF-MD) and body translation (**A**), the time from grabbing an object until it was deposited into the trolley (**B**), the head rotation (**C**) and the total number of grabbing motions for the target item (**D**). The IVF-MD is the mean deviation of the integrated visual field.*[6] *Error bars indicate mean ± standard deviation for each of the 8 glaucoma patients (represented with their ID and colour, see **Supplementary Table 1**). The corresponding grand mean value for the control participants is shown as a triangle on the y-axis. Unfilled circles*

*show the task performance for each item the glaucoma patients collected. The black line signifies the standard major axis regression, with shaded regions indicating the slope's 95% confidence interval. Black text at the top signifies Pearson's correlation coefficient and significance value.*
